# Where and how can WASH work? Understanding limited impacts from a randomized control trial of water, sanitation, and hygiene interventions in a high burden setting

**DOI:** 10.1101/2024.12.03.24318379

**Authors:** Alicia N. M. Kraay, Andrew F. Brouwer, Mondal H. Zahid, Sammy Njenga, John M. Colford, Matthew C. Freeman, Joseph N.S. Eisenberg

## Abstract

**Background:** Despite their strong theoretical basis, water, sanitation, and hygiene (WASH) interventions have had inconsistent benefits on diarrhea in low- and middle-income settings. The WASH Benefits (WASH-B) Kenya randomized controlled trial evaluated a set of WASH interventions targeted at children under age 2 and found no effect on diarrheal prevalence.

**Objectives:** We explored whether and how changes to intervention and contextual factors could impact health gains as a means to inform future WASH interventions.

**Methods:** We implemented a compartmental transmission model with environmental pathways and water (W), sanitation (S), and hygiene (H) interventions fit to WASH-B Kenya trial data (n=11,856) using a Bayesian sampling approach. We developed counterfactual simulations to predict how a trial might perform given improved 1) local contextual factors (i.e., reduced transmission, and increased completeness of transmission pathways targeted) and/or 2) intervention factors (i.e., increased intervention efficacy, compliance, and community coverage). We considered change in intervention effectiveness in the counterfactual scenarios for each intervention alone and in combination (WSH). We determined what combinations would be needed to achieve 50% reduction in child diarrhea compared to the control arm.

**Results:** We found that high diarrheal prevalence in the WASH-B Kenya trial was likely the primary reason for ineffectiveness. While none of the tested counterfactual factors greatly impacted intervention effectiveness in isolation, we estimated that 50% reduction in child diarrhea in the combined WSH treatment arm could be achieved through a combination of substantial intervention improvements (i.e., 50% intervention efficacy, 100% compliance, and 60% community coverage), but could not be attained for the single intervention arms. With improvements to contextual factors (consistent, 7.5% diarrheal prevalence, 50% increase in completeness) coupled with more modest increases in intervention factors (i.e., 50% efficacy and 100% compliance but only 20% community coverage), could achieve a 50% reduction in diarrhea in the combined WSH arm.

**Conclusions:** In settings with high enteric pathogen prevalence, WASH interventions must be used by a substantial fraction the population and block all main transmission routes to achieve substantial reductions in diarrheal disease burden, including those over age 2. The WASH interventions and targeting strategy for the WASH-B Kenya trial were unlikely to appreciably reduce diarrheal disease because of the high burden. In settings with more modest transmission, there are intervention factor targets that could result in measurable reductions in diarrhea. Application of this simulation-based approach could inform WASH policies and programs, as well as the design of future trials.

## Background

Enteric pathogens are a substantial contributor to under-5 mortality in low- and middle-income countries and are responsible for about 15% of all deaths globally among children under five years of age (1,2). Over the past decade, substantial progress has been made towards reducing mortality associated with diarrheal disease, largely attributed to rotavirus vaccination, reduced stunting, and improved access to treatment with oral rehydration solution, and, more recently, zinc supplementation (3). Yet, control of environmental exposure to diarrheal pathogens remains an important public health tool.

Water, sanitation, and hygiene (WASH) conditions and associated behaviors are thought to be responsible for nearly 70% of the global burden of diarrheal disease (2). Improved WASH can greatly reduce morbidity and mortality related to diarrheal disease. WASH interventions have a strong mechanistic basis and can lower exposure to enteric pathogens both through person-to-person contact and through the environment. While observational studies in low-income settings have found protective associations between WASH interventions and diarrheal disease risk, effect sizes have varied widely, potentially due to confounding factors (4,5). The WASH Benefits (WASH-B) studies in Kenya and Bangladesh were designed to robustly evaluate the impact of WASH interventions on diarrhea and child growth using a randomized control trial (6). While some of the interventions did reduce diarrheal prevalence in Bangladesh (7), particularly during the rainy season (8), no reduction diarrhea was seen in Kenya in any arm (9). A related trial in Zimbabwe conducted at about the same time also failed to find impact of WASH on diarrhea (10). These findings caused considerable discussion in the literature about the need for more transformational WASH interventions to greatly reduce the fecal contamination in the household environment (4,11,12). What it would take – technologically, behaviorally, programmatically - to achieve sufficient reduction in fecal pathogen exposure, especially for young children, remains an open question.

While randomized control trials are generally considered the gold standard for evaluating disease risk, results from individual trials may not be generalizable as the intervention effectiveness will likely differ across locations (13). Indeed, settings vary considerably in terms of their baseline disease burden, level of preexisting WASH infrastructure, and in the specific pathogens that are circulating, all of which can impact generalizability of any given RCT. Importantly, the causal attribution in RCTs are only as valid as the interventions delivered in this given context. In a recent study, Brouwer et al. investigated how impacts of the WASH-B trial in Bangladesh might have been different if there were changes to intervention and contextual factors (14). In this paper, we apply this mechanistic framework to the WASH-B Kenya trial, with the aim to both explain the lack of intervention effectiveness in the original trial and provide insight that can be used to improve the effectiveness of future WASH interventions.

## Methods

### Overview

In this study, we aimed to assess potential mechanistic reasons for the null results in the WASH-B Kenya trial and to identify opportunities to improve future public health interventions to maximize health gains. Specifically, we are interested in: 1) What set of interventions are effective in reducing transmission? and 2) What types of settings are likely to be most effective for WASH practitioners focus on?

### Kenya WASH B study data

The WASH Benefits Kenya study has previously been described in detail (6,9). In brief, 8,246 pregnant women in 702 clusters of one to three neighboring villages in Western Kenya having at least six pregnant women each were enrolled in the study, and their children under age 3 at baseline were then followed over a two-year follow-up period. At baseline, each cluster was randomized to receive either one of two control conditions (with or without community health promotor visits; labeled active and passive respectively), water treatment with chlorine (W), sanitation slab with lid and also received a sani-scoop to enable safe disposal of child feces (S), handwashing with soap and water (H), or nutritional supplement (N) interventions. There was also a combined water, sanitation and handwashing arm (WSH) and a water, sanitation, hygiene and nutrition arm (WSH-N). Data on adherence to the intervention and caregiver-reported all-cause diarrhea over the past 7 days in each child was collected at baseline, year 1, and year 2 of follow up. In this analysis, we used compliance and diarrheal assessments from all three time points and arm assignment to fit our transmission model. We excluded children with negative ages (n=132), children with missing diarrhea information (n=850), the passive control group (n=450), and those missing compliance information (n=4,446), leaving a total of 11,856 observations from 622 clusters, 67% of the original sample. The passive control group was excluded because use of water, sanitation, hygiene, and nutrition indicators were only measured at baseline and thus could not be accounted for by the model. In this analysis, we only used data that were publicly available at: https://osf.io/tprw2/.

We considered how intervention factors under the control of the investigator (called intervention factors) would need to be adjusted to deliver health gains and how these factors interact with features of the local context (contextual factors), showing when and where these gains might be achievable. We considered the following factors that could influence intervention effectiveness.

#### Intervention factors

- *Community coverage:* Because only households with pregnant women were enrolled in the RCT, we account for the fraction of all households in each cluster that were enrolled in the trial.
- *Intervention efficacy*: common WASH interventions generally reduce but do not eliminate transmission risk along a given transmission pathway, so we account for the degree to which the intervention, if used, reduces exposure to pathogens.
- *Compliance:* not all individuals randomized to receive an intervention received it or consistently use it, so we account for the adherence of individuals in the trial to their assigned intervention (and the fidelity of the study in providing the intervention as planned).

#### Contextual factors

- *Transmission intensity:* The baseline prevalence of diarrhea varied over time and across the intervention arms. We accounted for these differences to assess how baseline disease transmission impacts intervention effectiveness.
- *Preexisting WASH infrastructure:* Some individuals in each arm had access to improved WASH infrastructure prior to the start of the study. We account for this baseline protection when calculating effectiveness of each intervention arm.
- *Intervenable fraction:* not all transmission pathways are interrupted by WASH interventions, and different pathogens will exploit different transmission pathways to different degrees. For example, water treatment does not reduce exposure to pathogens transmitted through fomites. We thus account for how much transmission occurs along pathways targeted by the intervention and how much transmission even a perfect intervention would not be able to target. In our counterfactual simulations, we vary the intervenable fraction by 1) keeping the fitted basic reproduction number constant over time and 2) varying the fraction of transmission that is not intervenable.

To assess the importance of each intervention or contextual factor to the WASH-B Kenya trial results, we fit our previously developed transmission model to the WASH-B Kenya data for 7 arms simultaneously (active control, water treatment, sanitation, handwashing, nutrition, combined water, sanitation and hygiene and combined water, sanitation, hygiene and nutrition), and conducted counterfactual simulations with the fitted model to assess potential impacts by intervention arm given improvements in contextual factors, intervention factors, or both. We compared these counterfactual simulations to the original fitted trial scenario, and the difference between these arms provides insight into the contribution of each factor to the original trial outcomes.

### Transmission model and fitting

Our transmission model is a compartmental infectious disease transmission model that also tracks pathogen concentration in one or more local environmental pathways. This model has been described previously (14,15), and a reproduction of the corresponding model diagram is shown in Figure S1. In brief, the model divides each cluster into susceptible (*S*) and infectious (*I)* compartments and tracks disease prevalence for individuals with attenuated exposure from their WASH interventions (either due to baseline WASH infrastructure or due to the intervention rollout), hereafter called the ‘reduced exposure population,’ separately from the normal exposure population. Only those who are adhering to the intervention (ρ) are modeled as being protected; those who receive but do not adhere to the intervention have normal levels of exposure. Because we are modeling all-cause diarrhea, we assume that after recovery (γ) infected children become fully susceptible again. Infected individuals shed pathogens into the environmental compartments (*E_w_*, *E_f_*, *E_o_*) at rate α_j_, which are shared between all households in a cluster. Pathogens decay in the environment at rate *ξ_j_*. We model separate environmental compartments for the different transmission pathways, including water (*E_w_*), fomites/hands (*E_f_*), and other pathways (*E_o_*). Susceptible individuals can become infected (at rate β_j_) through contact with the contaminated environments. From steady-state data provided by the trial, parameters α, *ξ*, β, and y are not individually identifiable, but the pathway-specific basic reproduction numbers (in the absence of intervention), defined below, are.

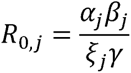

Depending on their level of WASH infrastructure, the reduced exposure population may have reduced shedding into the environment, reduced susceptibility to infection or contact with the environment, or both. The efficacy of each intervention (φ) is modeled uniquely based on which of these mechanisms is/are used. We also account for the fact that the amount of protection conferred by preexisting conditions may be different from that obtained by the intervention due to the lack of regular visits from health promoters to encourage continued use. Specifically, water treatment reduces transmission to households from the community water supply (*φ_βW_,W*). Sanitation is modeled as reducing shedding into the water environment and its efficacy is different for intervention and preexisting conditions (*φ_αw,S_*, *φ̄_αw,S_*). Handwashing reduces transmission from fomites and efficacy is different for intervention and preexisting conditions (*φ_βf_,H, φ̄_βf_,H*). Nutrition reduces transmission from all pathways, including the ‘other’ pathway (*φ_βN_,*).

We assumed that all study households in each cluster received the cluster-randomized intervention, but non-study households within the same cluster do not, although they may have some protection from preexisting WASH conditions within their own household. We assumed that the distribution of preexisting WASH conditions for non-study households in each arm was the same as the control arm at baseline. Because all interventions may be used in each cluster due to preexisting WASH conditions, regardless of the randomized intervention, we modeled adherence to each intervention separately in each cluster. Thus, we modeled the fraction of the population having protection from water treatment, sanitation, handwashing, nutrition and their various combinations for each arm. Based on this structure, our model has 24=16 infection compartments for each arm, or 32 compartments total (one for each S/I state), in addition to the three environmental compartments (see supplemental materials for model equations).

The transmission model described above is solved for equilibrium diarrhea prevalence (π = ∑*Ii*) in each of the 16 infection compartments in each cluster using a hybrid sampling- resampling algorithm using the nleqslv package in R as described in (14). In summary, we fit the following 18 model parameters to baseline diarrhea prevalence and relative risk data from the Kenya RCT: i) the basic reproduction number (R0), ii) two parameters that partition the overall reproduction number into the water (R0,*w*), fomite & hands (R0,*f*), and other pathways (R0,*o*), iii) eight scalar parameters that account for the relative transmissibility of each arm compared with the control group and differences between baseline, midline, and endline transmission, iv) community coverage, ω, and v) six overall efficacy parameters (φ), as described above. We randomly sampled 100,000 parameter combinations within the credible interval from each parameter using Sobol sampling and calculated the negative log likelihood for each parameter set. We then resampled, with replacement, from these 100,000 parameter combinations with a probability of sampling proportional to the negative log likelihood for each set, obtaining 3,110 distinct parameter combinations that were used in our final samples. The effectiveness (ε) of the intervention for each parameter set, *k*, was defined based on the relative prevalence of diarrhea compared with the control group 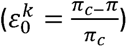. Fitted parameter values for the original trial scenario are shown in Table 1.

**Table 1.** Parameters used in the SISE-RCT transmission model. Fitted parameters were assumed to follow a uniform distribution, whose credible range is given in parentheses. The exception was *R*0,*F*, which was assumed to a uniform proportion of the quantity R0 – R0,*W* . In addition to the parameters shown below, we also fit the relative reproduction numbers of midline/endline compared with baseline and relative reproduction number by arm to adjust for arm-specific and time-varying confounding variables. Median values for fitted parameters are shown except for the parameters with an asterisk related to the component reproduction numbers (mean is shown instead). For the component reproduction numbers, the mean is shown instead due to the right-skewed shape of the component distributions for the water and fomite reproduction numbers (see Figure S2).

| Parameter | Definition | Value (credible range) | Source |
| --- | --- | --- | --- |
| **Fixed parameters** |  |  |  |
| $\pi_0$ | Baseline diarrheal prevalence | 0.158 | Data |
| $\rho_0$ | Proportion with baseline WASH conditions | W=0.019, S=0.135, H=0.054, WH=0.001, WS=0.008, HS=0.012, None=0.770 | Data |

| Compliance in treatment arm (average of midline and endline) |  |  | Data |
| --- | --- | --- | --- |
| $\rho_{1.5,W}$ | **W** Water treatment | 0.296 | |
| $\rho_{1.5,S}$ | **S** Sanitation | 0.815 | |
| $\rho_{1.5,H}$ | **H** Handwashing | 0.460 | |
| $\rho_{1.5,N}$ | **N** Nutrition | 0.516 | |
| $\rho_{1.5,WSH}$ | **WSH** | W=0.297, S=0.843, H=0.443 | |
| $\rho_{1.5,WSHN}$ | **WSH-N** | W=0.285, S=0.836, H=0.432, N=0.485 | |
| **Fitted parameters** |  |  |  |
| $R_0$ | Transmission potential | 1.19 (0, 1.65) | Fitted |
| $R_{0,w}/R_0$ | water | 0.21* | Fitted |
| $R_{0,f}/R_0$ | fomites | 0.25* | Fitted |
| $R_{0,o}/R_0$ | other | 0.54* | Fitted |
| $\omega$ | Community coverage fraction | 0.086 (0, 0.2) | Fitted |
| **Efficacy** |  |  | Fitted |
| $1 - \varphi_{\beta_W,W}$ | Efficacy of water at reducing transmission | 0.155 (0, 1) | |
| $1 - \varphi_{\alpha_W,S}$ | Efficacy of sanitation intervention at reducing shedding into $E_w$ | 0.421 (0, 1) | |
| $1 - \bar{\varphi}_{\alpha_W,S}$ | Efficacy of preexisting sanitation at reducing shedding into $E_w$ | 0.189 (0, 1) | |
| $1 - \varphi_{\beta_f,H}$ | Efficacy of handwashing intervention at reducing transmission from $E_f$ | 0.252 (0, 1) | |
| $1 - \bar{\varphi}_{\beta_f,H}$ | Efficacy of preexisting handwashing at reducing transmission from fomites $E_f$ | 0.093 (0, 1) | |
| $1 - \varphi_{\beta,N}$ | Efficacy of the nutrition intervention at reducing transmission (from all $E$ compartments) | 0.033 (0, 1) | |
| *Mean value for the ratios rather than median is shown so that the ratios of component R0 values add to 1 (see SI for details). |  |  |  |

### Counterfactual simulations

The goal of our counterfactual simulations was not to explore the full parameter space of all combined counterfactuals, but to identify combinations of interventions/scenarios that could produce at least a 50% reduction in diarrhea prevalence. The scenarios considered are shown in Table S1. For all scenarios, we ran simulations for the final 3,110 distinct resampled parameter combinations described above, and we summarized results by the median intervention effectiveness across the 100,000 resampled for each scenario. We reported the median of the difference between effectiveness in the counterfactual simulation and the effectiveness simulated in the original trial, namely the median absolute change in intervention effectiveness (*ε*- *ε*_0_).

We considered two general types of counterfactual scenarios. Changes to intervention factors are meant to represent changes that are, to some degree, under the control of the interventionist/researcher. In contrast, changes to contextual factors are not achievable through standard WASH interventions alone but instead reflect how a trial might perform if researchers were able to partner with other organizations to affect change in the underlying transmission system and/or if conducted in a different location with the specified changes in contextual factors. To explore these changes, we first explored how changing each intervention or contextual factor, one at a time, impacted the simulated effectiveness of the trial. Second, we explored the impact of combined changes to intervention factors, contextual factors, or both, might influence obtained effectiveness. We considered scenarios changing all contextual factors jointly, all intervention factors jointly, and whether or not all changes were needed simultaneously to achieve the 50% target reduction (see SI for intermediate scenarios). The counterfactuals considered in our model are a subset of those implemented by Brouwer et al, which have been defined previously (14). Due to our focus on WASH interventions, we did not include the N or WSH-N arms in counterfactual simulations but did use these arms to fit the original scenario and illustrate model performance.

We targeted a 50% reduction in diarrheal disease period prevalence to inform intervention targets that would be relevant to public health policy. In 2013, UNICEF and the World Health Organization set a global goal of ending preventable deaths from diarrhea, defined as 1 death per 1,000 live births from diarrhea each year (16). In the GEMS study conducted at about the same time as WASH -Benefits Kenya, diarrhea mortality was about 3.3 deaths per 1,000 (3). Thus, achieving the goal of ending preventable diarrhea deaths for Kenya corresponds to a roughly 67% reduction in diarrheal deaths. For simplicity and due to the fact that we modeled prevalence and not deaths from disease (and given improvements in treatment, fewer cases need to be averted to prevent a single death), we apply a less stringent “50% reduction” benchmark for all cause diarrhea incidence in our simulations.

## Results

Our simulated prevalence of diarrheal disease by arm at midline/endline and closely matched the data from the trial (Figure S3). The median intervention effectiveness values corresponding to these fits were W=3.0%, S=-2.4%, H=8.3%, WSH=1.4%, N=2.1%, WSHN=-1.7%, all of which fell within the 95% confidence intervals of the published original statistical analysis (9).

We considered how increasing intervention efficacy (to 50% or 100%), improving compliance (to 100%), reducing baseline diarrhea by half, having stable transmission intensity over the study period (data suggested an increase in transmission, so we modeled a scenario where this increase in transmission over time did not occur), and reducing the strength of the ‘other’ pathway might improve predicted effectiveness for each arm of the trial. In general, changing single intervention or contextual factors did not appreciably change the simulated effectiveness values for the W, S, H, or WSH arms (Table S2, Figure 1) and fell far short of the goal of reducing diarrhea by 50%. The strongest gains for single interventions were achieved in the WSH arm with 100% efficacy of all three interventions, with an expected intervention effectiveness of about 24%.

**Figure 1.**
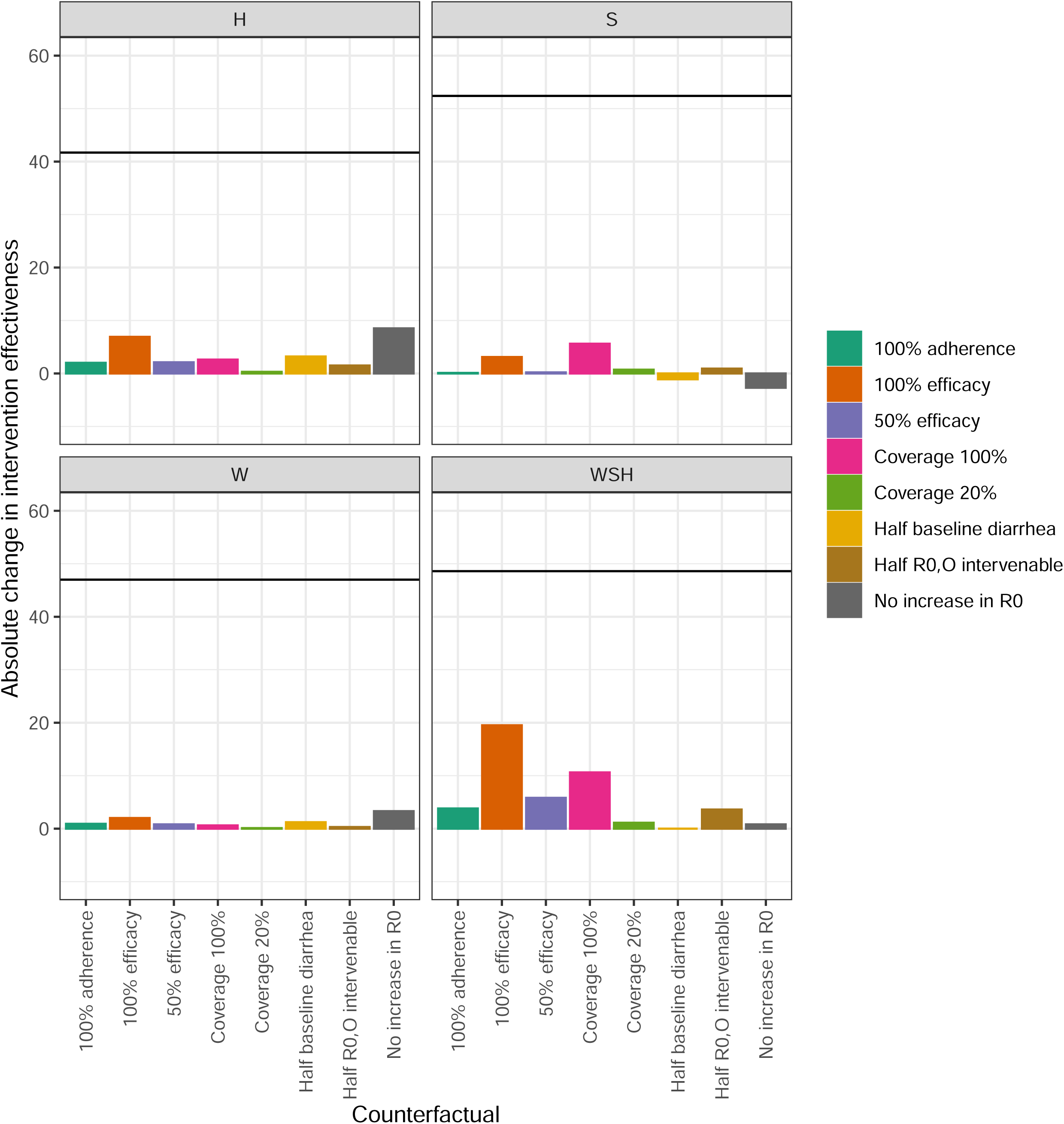
Simulated absolute change in intervention effectiveness (%) by arm for individual changes in counterfactuals. The solid line shows the needed absolute change corresponding to the 50% efficacy target for each arm. Bars show changes to intervention factors (efficacy, adherence, coverage) and contextual factors (half baseline diarrhea, less other transmission, no change in R0 over time). W=water, S=sanitation, H=handwashing, WSH=water, sanitation, and handwashing.

We also considered the impact of changing intervention factors jointly with varying levels of increased intervention community coverage (note that the original trial focused on only intervening in multi-family compounds where children under age 2 resided, given the focus on measuring health benefits among this target age group that spend relatively little time outside their home environment; at the time of enrollment, only 9% of the community was eligible for enrollment in the trial on the basis of having a pregnant woman). In simulations with moderate increases in intervention factors where adherence was set to 100% and efficacy was set to a moderate value of 50%, we were able to achieve intervention effectiveness greater than 50% in the WSH arm when coverage was above 60% (Figure 2, effectiveness at 60% coverage = 47.1%). In contrast, even with 100% community coverage, the increase in compliance and intervention efficacy could not deliver the desired 50% reduction in the W (effectiveness = 15.8%), S (effectiveness =8.7%), and H arms (effectiveness =30.3%). The impact of increasing coverage was approximately linear and small in the W, S, and H arms but had a much larger and moderately increasing trend in the joint WSH arm, with increasing coverage providing a stronger benefit at higher levels of coverage. For example, in the W arm, the absolute increase in intervention effectiveness when increasing community coverage from 20 to 40% was 1.9% and 2.1% when increasing community coverage from 80 to 100%. In the WSH arm, increasing community coverage from 20 to 40% provided an 11.4% absolute increase in intervention effectiveness, whereas increasing from 80 to 100% provided a 15.2% absolute increase.

**Figure 2.**
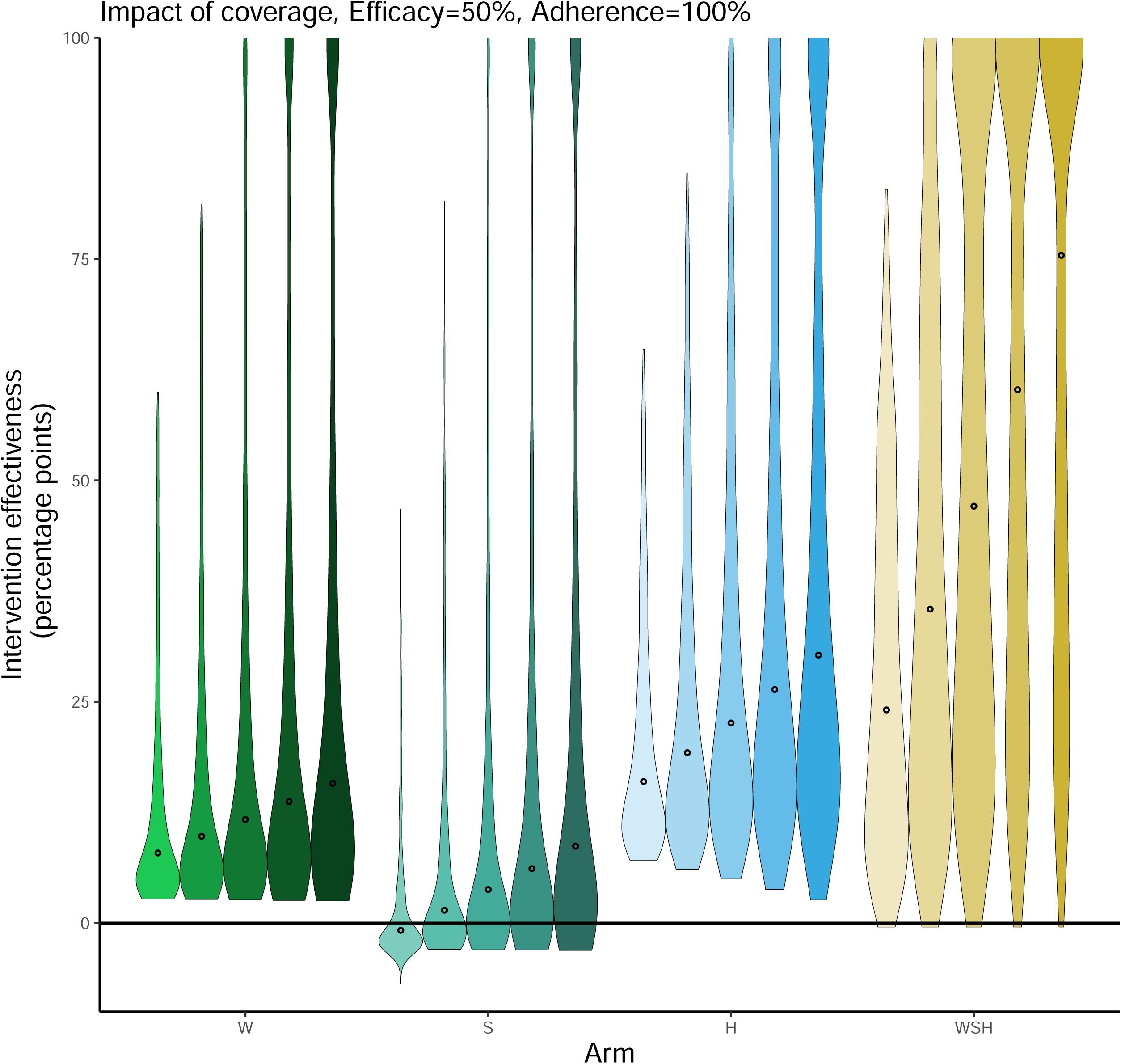
Simulated intervention effectiveness given 50% efficacy and 100% compliance by quintile of community coverage (20%, 40%, 60%, 80%, and 100%). Points show the median values across 100,000 simulations with violin plots showing the distribution. W=water, S=sanitation, H=handwashing, WSH=water, sanitation, and handwashing

In addition, we simulated what impacts might have been if the trial were repeated in a different location with more favorable contextual factors (50% less ‘other’ transmission, constant transmission intensity, half the baseline diarrheal prevalence) with or without the same changes in intervention factors (Figure 3) and with coverage fixed at 20%. We found that improving contextual factors yielded improved effectiveness values near 50% in the H arm (ε=41.3%), but intervention effectiveness remained low in the W (ε=14.8%), S (ε=-7.4%), and WSH arms (ε=14.2%). However, when contextual factors were changed with moderate improvements in intervention factors, predicted intervention effectiveness was >50% in the W (ε=52.9%), H (ε=100%), and WSH arms (ε=100%). While below 50%, effectiveness in the S arm was also substantially improved (ε=19.1%).

**Figure 3.**
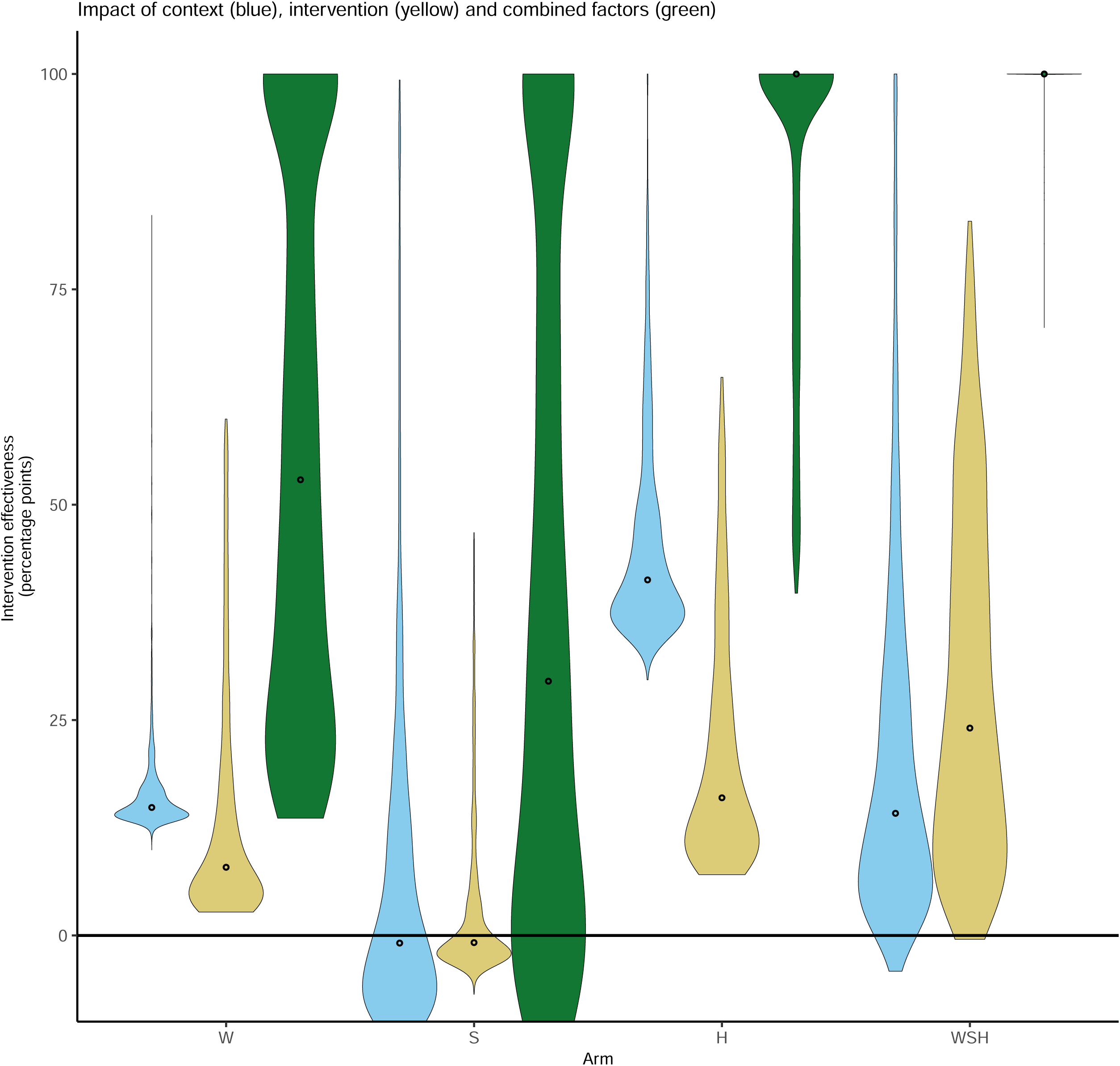
Combined counterfactuals. The expected effectiveness given contextual changes alone (50% less ‘other’ transmission, constant transmission intensity, half baseline diarrheal prevalence) is shown in blue, intervention changes alone (adherence=100%, coverage=20%, efficacy=50%) is shown in yellow and changes in both factors combined is shown in green.

When considering the WSH arm, changes in contextual factors needed to increase effectiveness over 50% were smaller, when compared to individual arms (Table S2). In particular, it was necessary that only two of the three considered improvements in contextual factors were met (50% less ‘other’ transmission, constant transmission intensity, half the baseline diarrheal prevalence) when combined with the same improvements in intervention factors. For more favorable transmission settings, when all three contextual factor conditions were met, less improvement in intervention factors (two out of three of 20% coverage, full compliance, or 50% efficacy) were needed for effectiveness in the WSH arm to exceed 50% (Table S2). In addition, if all three contextual factors were met and community coverage could be increased to 40%, predicted effectiveness of the WSH intervention arm exceeded 50% (*ε* = 60.7%) (Table S2).

## Discussion

In general, we find that understanding the contextual factors in a population and carefully considering intervention factor targets is important to realize desired reductions in disease burden. In context with high disease burden, standard WASH interventions alone are unlikely to be enough and multi-pronged approaches will likely be needed. Such interventions will need to target a broader range of disease transmission pathways and achieve high coverage. On the converse, reductions in local disease transmission pressure can make achieving these benefits more feasible.

Specifically, in our model simulation study, when WASH interventions were delivered alone we found that achieving the UNICEF target of 50% reduction in diarrhea prevalence could be achieved with a combination of higher efficacy interventions, higher intervention adherence, and targeting a larger fraction of the community for intervention, but the level of intervention required to achieve these gains in high burden settings may make such high effectiveness difficult to impossible to achieve in practice. Combining water, sanitation, and hygiene interventions – as is typically done programmatically - requires lower efficacy, compliance, and coverage to achieved the commensurate targets. A sequenced set of interventions that can additionally target contextual factors (baseline diarrheal disease, etc.) provides potentially a more sustainable transformative solution. For example, carefully planned infrastructure that accounts for multiple levels of risk is more resilient, and may remain more effective in the face of natural disasters like floods could help allow diarrheal disease incidence to remain more stable (17). In addition, fly control measures like reducing breeding sites could lower the transmission potential from other pathways that cannot be impacted as readily by WASH interventions (18). If reduction of diarrheal disease – or another infectious disease – is the goal, our findings point to the importance of understanding the context, and adjusting the intervention factors accordingly. Infection pressure is perhaps the most crucial contextual factor as high burden settings require much higher levels of efficiency, coverage, and compliance to be effective.

In our analysis, some of the intervention factor targets may be considered programmatically ambitious, while others may be more achievable. How achievable the efficacy for each intervention may depend on which pathogens are circulating and what other interventions are present in the community. For example, the 50% efficacy value is similar to the median estimated efficacy of the same water chlorination intervention in Bangladesh (14), suggesting that this target can be achieved in some low resource settings. It’s possible that treating water with chlorine can achieve up to 100% efficacy for removing *E. coli* pathogens from water (19). However, chlorination is not efficacious against some pathogens, including *Cryptosporidium* (20), and is less efficacious for some viruses (21). Handwashing has high efficacy for many pathogens but is known to not be as efficacious against rotavirus (22).

Handwashing with non-potable water can increase risk, suggesting that efficacy may be higher in settings where water treatment interventions are also implemented (23). We modeled sanitation as reducing the shedding of fecal material into drinking water sources, a major route of transmission for norovirus, rotavirus and cryptosporidium (24). However, sanitation may also reduce risks from other transmission pathways that are not explicitly modeled in this paper, such as transmission from flies (24). Thus, the achieved efficacy for WASH interventions may depend on the pathogens in circulation and local contextual factors.

Achieving high levels of intervention coverage is essential to interrupt transmission in a high burden setting by changing intervention factors alone. WASH is typically delivered at a community, rather than household level, but even so, coverage improvements from WASH interventions have proven relatively modest. As an example, sanitation interventions have been shown to increase coverage by 14 percentage points on average (25). Coverage targets for interventions must be much more ambitious to deliver on promised health gains among the most vulnerable. Future trials of WASH interventions should ensure that intervention coverage is high, even if trial enrollment is only targeted to measure the health effects of children under five, as was done in field trials of at-scale sanitation interventions in Mali and Odisha, India (26,27).

Perfect compliance is likely unrealistically high for some interventions. While intervention compliance was relatively high for the sanitation arm (82-84%) in the Kenya trial, compliance was substantially lower for water treatment (∼30%) and handwashing (43-46%). However, novel technologies that require no user effort have been developed for piped water treatment and have been shown to achieve strong benefits (23% reduction in child diarrhea), even with imperfect efficacy of around 77% (based on the difference in E. coli detection between intervention and control arms) presumably due to the lack of user effort required (10,28). For water systems without piped water, like the Kenya study site would require other innovations (6). The goal of WASH interventions is to increase effectiveness, which may be easier to achieve with high compliance, even if efficacy is slightly lower (29,30). Thus, reducing user burden could have major implications for increasing intervention effectiveness by increasing adherence.

WASH interventions can deliver substantial benefits, but our analysis suggests that achieving these benefits will require not only targeting multiple WASH interventions but also adding to this interventions that target contextual factors, particularly in high burden settings. Integrating interventions that target other pathways of diarrheal prevention and treatment may be especially important in high burden settings. Additional evidence from the Kenya study region has provided some suggestion that these contextual changes may already be occurring. For example, in the recent VIDA study, diarrheal mortality in western Kenya site declined by 86% between 2008 (GEMS) and 2018 (VIDA). The largest factors estimated as contributing to this decline were increased access to zinc, reductions in wasting prevalence, and expanded rotavirus vaccination (3). Given the noted impact of rotavirus vaccination on local transmission dynamics in Kenya, the variety of other enteric vaccines that are currently under development (i.e., shigella, norovirus) may also help reduce baseline burden (31,32), creating an environment in which WASH can work.

Our study has several limitations. Notably, we used an all-cause diarrhea transmission model simulated at equilibrium. However, the impact of WASH interventions is likely to be pathogen specific and the pathogens that are dominant may vary by location (33). In addition to spatial variation, dominant pathogens as well as the transmission intensity are likely to vary by season (33,34), whereas our model averages across the cases that occurred over each year of follow up. Such seasonal variation was shown to be important in the WASH Benefits- Bangladesh study, where impacts of WASH interventions were substantially greater during rainy seasons (8), in contrast to the Kenya study where there was no evidence of seasonality in diarrhea prevalence. While we include multiple transmission pathways explicitly in our model, we are not able to account for all potential pathways of transmission, including fly transmission (35) or transmission from contact with animals (36). These additional pathways are captured by the ‘other’ pathway in our model, but WASH interventions may not work uniformly against these other pathways. Nevertheless, we feel that this model represents an important step forward in understanding WASH impacts in high burden settings, and our framework can be expanded in future work to capture these and other nuances.

## Conclusions

Water and sanitation are human rights, and are fundamental to human development and economic growth (37). There are further many public health justifications for improving water, sanitation, and hygiene, including mitigating the burden of infectious disease; improving child growth; addressing privacy, dignity, and well-being. One of the key drivers for improving WASH is the potential to mitigate diarrheal disease burden in high burden settings among the most vulnerable populations. Our study underscores the importance of understanding the contextual factors in a population and carefully considering intervention factor targets to ensure achievement of anticipated and policy relevant reductions in disease burden. Specifically, high baseline disease burden can make succeeding with standard WASH interventions more difficult. In such contexts, WASH interventions that are multi-faceted that are both more efficient, delivered at higher coverage, and block a larger number of transmission pathways are needed. As baseline burden declines, the full potential of WASH interventions may become easier to realize. Our findings, though derived from one specific study in Kenya, have broad implications for WASH programs, policies, and intervention field trials, not just for diarrheal diseases, but for other infectious diseases and pathogens as well where WASH can play a crucial role.

## Data Availability

All data that are used in this manuscript are publicly available at: https://osf.io/tprw2/.

## Notes

### Competing Interest Statement

Conflicts of interest: ANMK's contributions were directly funded by the Bill & Melinda Gates Foundation and not as part of the foundation grant to the authors. All other authors declare no conflicts of interest.

### Funding Statement

This study was funded by the Bill & Melinda Gates Foundation (INV-005081).

### Author Declarations

In this analysis, we only used data that were publicly available at: https://osf.io/tprw2/.

