## Supplementary material for "Where and how can WASH work? Understanding limited impacts from a randomized control trial of water, sanitation, and hygiene interventions in a high burden setting": SI

**Supplemental Material**

**Figure S1.** Compartmental model diagram used in the simulations, as reproduced from (15).

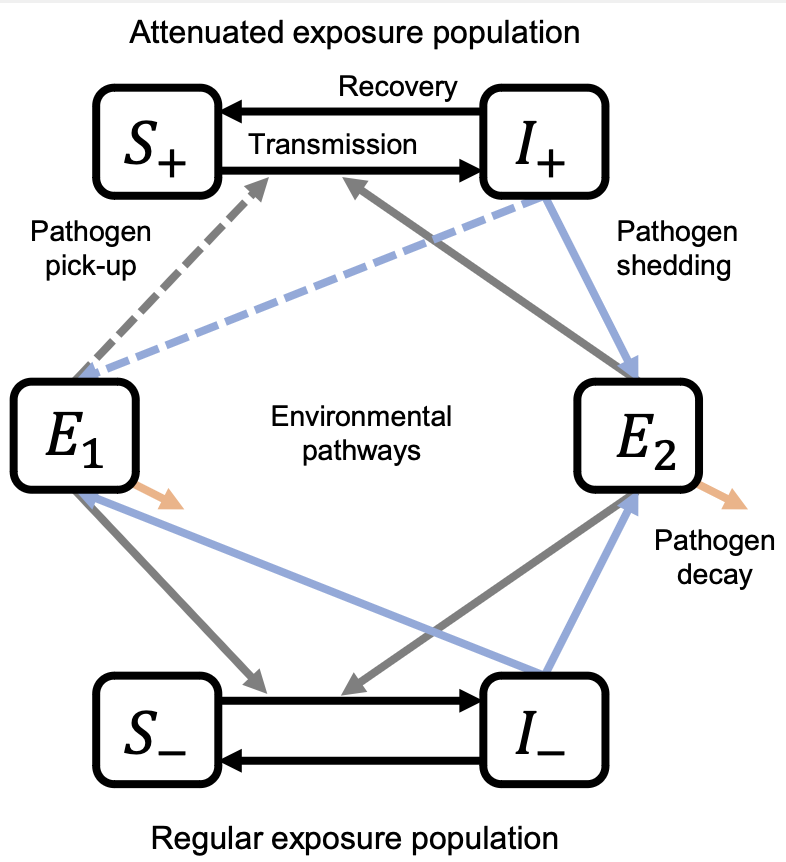

**Model equations**

Model equations for the I and E compartments are shown below. The corresponding Si compartments can be written as $\frac{{dS}_{i}}{dt}=-\frac{{dI}_{i}}{dt}$ for each subpopulation, i.

$$\frac{{dI}_{0}}{dt}=\left( \beta_{w}E_{w}+\beta_{f}E_{f}+\beta_{o}E_{o} \right)S_{0}-\gamma I_{0}$$

$$\frac{{dI}_{W}}{dt}=\left( {\varphi_{\beta_{w},W}\cdot\beta}_{w}E_{w}+\beta_{f}E_{f}+\beta_{o}E_{o} \right)S_{W}-\gamma I_{W}$$

$$\frac{{dI}_{S}}{dt}=\left( \beta_{w}E_{w}+\beta_{f}E_{f}+\beta_{o}E_{o} \right)S_{S}-\gamma I_{S}$$

$$\frac{{dI}_{H}}{dt}=\left( \beta_{w}E_{w}+{\varphi_{\beta_{f},H} \cdot\beta}_{f}E_{f}+\beta_{o}E_{o} \right)S_{H}-\gamma I_{H}$$

$$\frac{{dI}_{N}}{dt}=\varphi_{\beta,N}\left( \beta_{w}E_{w}+\beta_{f}E_{f}+\beta_{o}E_{o} \right)S_{N}-\gamma I_{N}$$

$$\frac{{dI}_{WS}}{dt}=\left( {\varphi_{\beta_{w},W}\cdot\beta}_{w}E_{w}+\beta_{f}E_{f}+\beta_{o}E_{o} \right)S_{WS}-\gamma I_{WS}$$

$$\frac{{dI}_{WH}}{dt}=\left( {\varphi_{\beta_{w},W}\cdot\beta}_{w}E_{w}+{\varphi_{\beta_{f},H} \cdot\beta}_{f}E_{f}+\beta_{o}E_{o} \right)S_{WH}-\gamma I_{WH}$$

$$\frac{{dI}_{WN}}{dt}=\varphi_{\beta,N}\left( {\varphi_{\beta_{w},W}\cdot\beta}_{w}E_{w}+\beta_{f}E_{f}+\beta_{o}E_{o} \right)S_{WN}-\gamma I_{WN}$$

$$\frac{{dI}_{SH}}{dt}=\left( \beta_{w}E_{w}+{\varphi_{\beta_{f},H} \cdot\beta}_{f}E_{f}+\beta_{o}E_{o} \right)S_{SH}-\gamma I_{SH}$$

$$\frac{{dI}_{SN}}{dt}=\varphi_{\beta,N}\left( \beta_{w}E_{w}+\beta_{f}E_{f}+\beta_{o}E_{o} \right)S_{SN}-\gamma I_{SN}$$

$$\frac{{dI}_{HN}}{dt}=\varphi_{\beta,N}\left( \beta_{w}E_{w}+\varphi_{\beta_{f},H} \cdot\beta_{f}E_{f}+\beta_{o}E_{o} \right)S_{HN}-\gamma I_{HN}$$

$$\frac{{dI}_{WSH}}{dt}=\left( {\varphi_{\beta_{w},W}\cdot\beta}_{w}E_{w}+{\varphi_{\beta_{f},H} \cdot\beta}_{f}E_{f}+\beta_{o}E_{o} \right)S_{WSH}-\gamma I_{WSH}$$

$$\frac{{dI}_{WSN}}{dt}=\varphi_{\beta,N}\left( {\varphi_{\beta_{w},W}\cdot\beta}_{w}E_{w}+\beta_{f}E_{f}+\beta_{o}E_{o} \right)S_{WSN}-\gamma I_{WSN}$$

$$\frac{{dI}_{WHN}}{dt}=\varphi_{\beta,N}\left( {\varphi_{\beta_{w},W}\cdot\beta}_{w}E_{w}+{\varphi_{\beta_{f},H} \cdot\beta}_{f}E_{f}+\beta_{o}E_{o} \right)S_{WHN}-\gamma I_{WHN}$$

$$\frac{{dI}_{SHN}}{dt}=\varphi_{\beta,N}\left( \beta_{w}E_{w}+{\varphi_{\beta_{f},H} \cdot\beta}_{f}E_{f}+\beta_{o}E_{o} \right)S_{SHN}-\gamma I_{SHN}$$

$$\frac{{dI}_{WSHN}}{dt}=\varphi_{\beta,N}\left( {\varphi_{\beta_{w},W}\cdot\beta}_{w}E_{w}+{\varphi_{\beta_{f},H} \cdot\beta}_{f}E_{f}+\beta_{o}E_{o} \right)S_{WSHN}-\gamma I_{WSHN}$$

$$\frac{{dE}_{w}}{dt}=\alpha_{w}\left( \sum_{S not in i} I_{i}+ \varphi_{\alpha,S} \sum_{S in i} I_{i} \right)-\xi_{w}E_{w}$$

$$\frac{{dE}_{f}}{dt}=\alpha_{f}\left( \sum_{i} I_{i} \right)-\xi_{f}E_{f}$$

$$\frac{{dE}_{o}}{dt}=\alpha_{o}\left( \sum_{i} I_{i} \right)-\xi_{o}E_{o}$$

**Pathway specific reproduction numbers**

The value of R_0_ was fitted for the model. To fit the pathway specific R_0_ values, we fit scalars R_adj,W_ and R_adj,H_ such that the pathway specific R_0_ values summed to the combined reproduction number. Specifically:

$$R_{0,W}=R_{0}\times R_{adj,W}$$

$$R_{0,H}=R_{0}\times(1-R_{adj,W})\times(R_{adj,H})$$

$$R_{0,other}=R_{0}\times(1-R_{adj,W})\times(1-R_{adj,H})$$

**Figure S2.** Reproduction numbers for a) all pathways, b) the water route, c) the fomite route, and d) the other route. For each panel, the mean is shown in blue and the median is shown in grey.
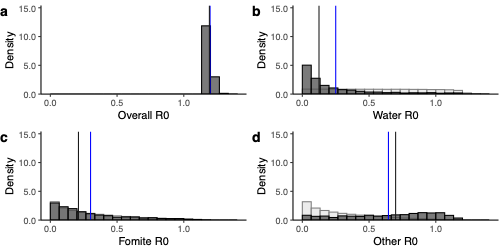

**Figure S3.** Simulated vs. measured prevalence of diarrheal disease by arm at baseline (red) and midline/endline combined (blue). Simulated values are shown as violin plots and data with confidence intervals are shown as points with error bars.

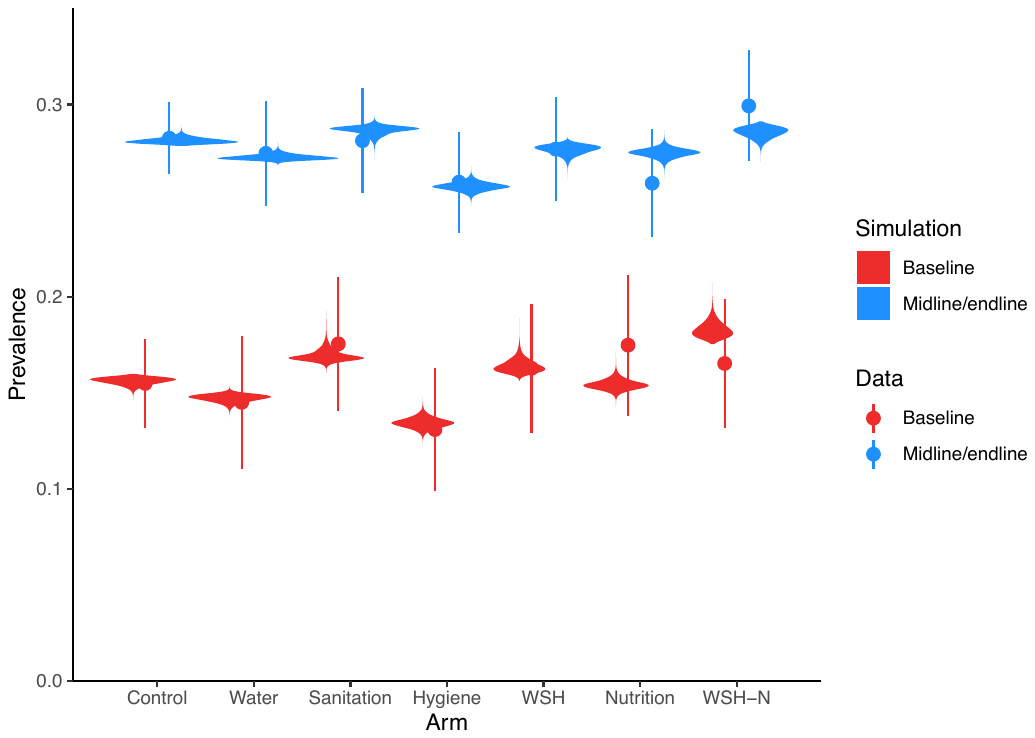

**Table S1.** Scenarios considered in main text.

| Scenario | Grouping |
| --- | --- |
| 50% less other transmission | Contextual |
| Constant transmission intensity | Contextual |
| Half baseline diarrheal prevalence | Contextual |
| 50% less other + constant transmission intensity + half baseline diarrhea | Contextual |
| 100% adherence | Intervention |
| 20% coverage | Intervention |
| 40% coverage | Intervention |
| 60% coverage | Intervention |
| 80% coverage | Intervention |
| 100% coverage | Intervention |
| 50% efficacy | Intervention |
| 100% efficacy | Intervention |
| 100% adherence + 20% coverage + 50% efficacy | Intervention |

**Table S2.** Counterfactual point estimates from counterfactual simulations for the combined water, sanitation, and hygiene (WSH) arm.

|  | *ε* | *Δε* |
| --- | --- | --- |
| ***Contextual Factors Only*** |  |  |
| Half baseline diarrhea + No increase in R0 + half other pathway intervenable | 14.2% | +12.3% |
| ***Intervention Factors Only*** |  |  |
| Full efficacy + Full adherence + full coverage | 100% | +98.6% |
| 50% efficacy + full adherence + full coverage | 75.4% | +74.3% |
| 50% efficacy + full adherence + 80% coverage | 60.2% | +58.6% |
| 50% efficacy + full adherence + 60% coverage | 47.1% | +45.5% |
| 50% efficacy + 80% coverage | 26.8% | +25.0% |
| 50% efficacy + full adherence | 18.4% | +16.8% |
| Full adherence + 80% coverage | 26.0% | +24.5% |
| ***Intervention + Contextual Factors*** |  |  |
| *All interventions* |  |  |
| 50% efficacy + full adherence + 20% coverage + half baseline diarrhea + no increase in R0 + half other pathway intervenable | 100% | +98.5% |
| *Stepwise removal of contextual factors (all intervention)* |  |  |
| 50% efficacy + full adherence + 20% coverage + half baseline diarrhea + no increase in R0 | 70.7% | +68.8% |
| 50% efficacy + full adherence + 20% coverage + half baseline diarrhea + half other pathway intervenable | 55.5% | +54.0% |
| 50% efficacy + full adherence + 20% coverage + no increase in R0 + half other pathway intervenable | 71.3% | +69.8% |
| 50% efficacy + full adherence + 20% coverage + no increase in R0 | 40.1% | +38.3% |
| 50% efficacy + full adherence + 20% coverage + half baseline diarrhea | 30.2% | +28.2% |
| 50% efficacy + full adherence + 20% coverage + half other pathway intervenable | 45.7% | +44.2% |
| *Stepwise removal of intervention factors (all context)* |  |  |
| 50% efficacy + full adherence + half baseline diarrhea + no increase in R0 + half other pathway intervenable | 79.1% | +77.2% |
| Full adherence + 20% coverage + no increase in R0 + half other pathway intervenable | 61.2% | +59.7% |
| 50% efficacy + 20% coverage + half baseline diarrhea + no increase in R0 + half other pathway intervenable | 73.1% | +71.7% |
| 50% efficacy + half baseline diarrhea + no increase in R0 + half other pathway intervenable | 46.9% | +45.2% |
| Full adherence + half baseline diarrhea + no increase in R0 + half other pathway intervenable | 29.8% | +27.8% |
| 20% coverage + half baseline diarrhea + no increase in R0 + half other pathway intervenable | 31.6% | +29.8% |
| 40% coverage + half baseline diarrhea + no increase in R0 + half other pathway intervenable | 60.7% | +59.3% |
| 60% coverage + half baseline diarrhea + no increase in R0 + half other pathway intervenable | 91.0% | +89.8% |
